# A COVID-19 transmission model informing medication development and supply chain needs

**DOI:** 10.1101/2020.11.23.20237404

**Authors:** Annabelle Lemenuel-Diot, Barry Clinch, Aeron C. Hurt, Paul Boutry, Johann Laurent, Mathias Leddin, Stefan Frings, Jean Eric Charoin

## Abstract

Accurate prediction of COVID-19 cases can optimize clinical trial recruitment, inform mitigation strategies and facilitate rapid medication development. Here we present a country-specific, modified Susceptible, Exposed, Infectious, Removed (SEIR) model of SARS-CoV-2 transmission using data from the Johns Hopkins University COVID-19 Dashboard. Inter-country differences in initial exposure, cultural/environmental factors, reporting requirements and stringency of mitigation strategies were incorporated. Asymptomatic patients and super-spreaders were also factored into our model. Using these data, our model estimated 65.8% of cases as asymptomatic; symptomatic and asymptomatic people were estimated to infect 2.12 and 5.83 other people, respectively. An estimated 9.55% of cases were super-spreaders with a 2.11-fold higher transmission rate than average. Our model estimated a mean maximum infection rate of 0.927 cases/day (inter-country range, 0.63–1.41) without mitigation strategies. Mitigation strategies with a stringency index value of ≥60% were estimated to be required to reduce the reproduction ratio below 1. It was predicted that cases over the next 2 months would differ between countries, with certain countries likely to experience an accelerated accumulation of cases. Together, results from our model can guide distribution of diagnostic tests, impact clinical trial development, support medication development and distribution and inform mitigation strategies to reduce COVID-19 spread.

**Key Findings:**

- Predicting COVID-19 cases can inform medication development and mitigation strategies
- We created a modified SEIR model of SARS-CoV-2 transmission
- We integrated asymptomatic cases, super-spreaders and hotspots that drive viral spread
- Mitigation strategies with a stringency index of ≥60% are required to reduce the RR below 1
- Some countries may experience an accelerated accumulation of cases in the coming months

## Introduction

As of 25 October 2020, the coronavirus disease 2019 (COVID-19) first reported in Wuhan, China, in December 2019 had resulted in 43 million confirmed cases globally, with infections continuing to spread [1]. This unprecedented pandemic has presented unique challenges for medical professionals, biomedical researchers, governmental and non-governmental organizations and members of the pharmaceutical industry, each of whom have shown an unwavering commitment to patient care and support [2, 3]. Specifically, the pharmaceutical industry has increased efforts to research, develop, register and make available solutions ranging from antivirals to treatments of complications of COVID-19 in record speed while also carefully managing supply lines and manufacturing sites for existing medications in high demand for the general management of patients with COVID-19 [4].

To support clinical trial recruitment, medication development, medication supply and distribution strategies, it is vital for the pharmaceutical industry, national and multi-national organizations, governments and non-governmental organizations to understand the epidemiological concept of virus transmission, the patterns and implications of severe acute respiratory syndrome coronavirus 2 (SARS-CoV-2) viral spread and the impact of different non-pharmaceutical interventions (NPIs) proposed as mitigation strategies on local, national and international levels [5, 6]. Specifically, the ability to accurately project the number of expected cases in each country over time could assist in selecting clinical trial sites with good potential for rapid patient recruitment and medication development. Furthermore, this information could guide the fair and equitable distribution of diagnostic tests, treatment options and vaccines.

The basic reproduction number (R_0_) for SARS-CoV-2 infection before the implementation of mitigation strategies is estimated to range from approximately 2.0 to 3.6 [7-12], with a high risk for transmission because of high numbers of asymptomatic subjects and emerging clusters [13-15]. Transmission models based on real-world epidemiological data are important tools for understanding the dynamics of SARS-CoV-2 transmission and can be useful to guide mitigation strategies and policy decisions designed to assist patients with COVID-19 and reduce disease spread [9, 16-21].

Mitigation strategies with NPIs have been effective in helping to curb the spread of SARS-CoV-2 and in reducing the reproduction ratio (RR) [12, 22], though these are likely hindered by the relatively high proportion of asymptomatic cases of COVID-19 [15, 23, 24]. Although the magnitude of infectiousness in asymptomatic patients (i.e. when the infector has no symptoms throughout the course of the disease) is difficult to quantify, these cases are expected to heavily impact transmission dynamics [25, 26]. Indeed, non–peer-reviewed mathematical modelling studies highlight the importance of accounting for asymptomatic persons when describing transmission dynamics [27, 28]. As such, this analysis could support public health considerations and suggest, by quantifying their contributions, that asymptomatic persons might be major drivers of the COVID-19 pandemic. Unless asymptomatic persons happen to get tested, they may continue to socialize and work during the entire infectious period, in contrast to situations in which the vast majority of people are symptomatic and can be identified without testing and for whom rapid quarantine is possible.

Although some components of viral transmission (e.g. proportions of asymptomatic and “super-spreader” cases, duration of latency, pre-symptomatic infectious and post-symptomatic infectious periods and duration of infectiousness) are likely to be consistent across countries, other components affecting patterns of viral spread are expected to differ between countries, further complicating potential models of viral transmission. Further, cases of COVID-19 are not uniformly distributed within a country; rather, they are primarily located in “hotspots” of various sizes that, without mitigation, merge and grow, potentially including the entire population [29-31]. A proportion of super-spreaders has also been reported in the COVID-19 population, and a limitation of classical models is the use of mean parameter values across the population, even though different persons may have different disease characteristics (e.g. viral load, infection rate, duration of symptoms). Additional proposed modelling approaches could account for super-spreader profiles by differentiating this type of case and estimating specific transmission characteristics of super-spreaders.

Although several epidemiological transmission models exist, our model is the first to clearly quantify the effect of NPIs on COVID-19 transmission in individual countries while also accounting for the expected contribution of asymptomatic cases to COVID-19 transmission and differentiating potential super-spreaders. Given that these components are crucial for robust, country-specific projections, we aimed to implement a modified Susceptible, Exposed, Infectious, Removed (SEIR) model of SARS-CoV-2 transmission incorporating those components, with the objectives of supporting the development of medications, optimizing clinical trial recruitment and facilitating a fast-to-market strategy for medications that have the potential to reduce symptoms and complications in patients with COVID-19.

## Methods

### Data sources

Real-world epidemiological data were obtained from the Johns Hopkins University COVID-19 Center for Systems Science and Engineering COVID-19 Dashboard on 31 August 2020 [1]. This includes data from national and state government health departments and local media reports. In addition, country-level mitigation data from the Coronavirus Government Response Tracker, collected and validated by Oxford University [32], were used to investigate the potential mitigation impact of NPIs on the transmission of SARS-CoV-2, with the objective of building country-specific quantitative relationships between NPIs and transmission model parameters.

### Model development

Development of the initial modified SEIR model was based on the susceptible population (S), exposed patients not yet infectious (E), infected infectious patients who are asymptomatic (Ia), infected infectious patients (I), recovered patients (R) and death (D) (Fig. 1). Additional components, detailed below, were added to account for reporting rates of individual countries, incorporate hotspots and emerging clusters, include asymptomatic and super-spreader profiles and evaluate the impact of various mitigation strategies on transmission rate. The model was used to estimate the expected total number of symptomatic and asymptomatic cases regardless of whether they were reported.

**Figure 1.**
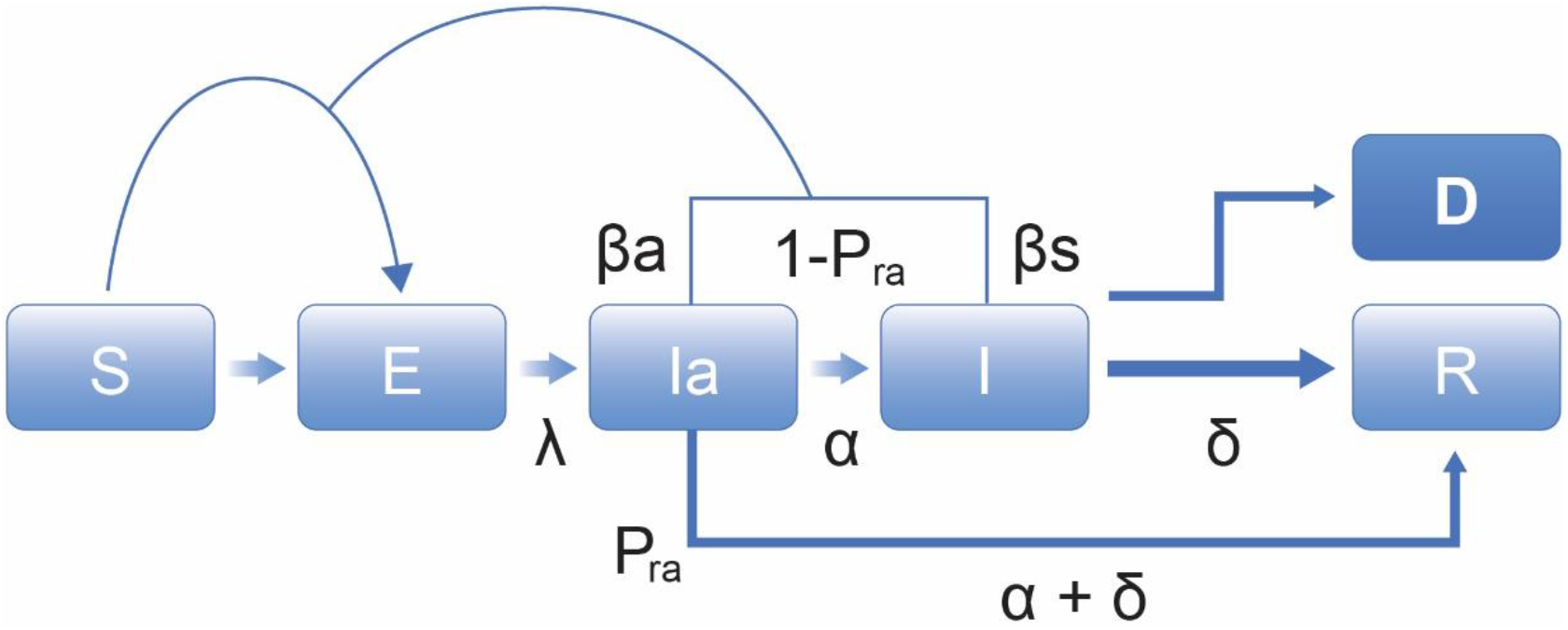
Initial COVID-19 transmission model. S, susceptible population; E, exposed patients not yet infectious; Ia, infected infectious patients who are asymptomatic; I, infected infectious patients; R, recovered; D, death. λ, median incubation period; α, start of infectious period; δ, duration of infectious period.

To account for regional differences in initial exposure, we started with a set time of 1 January 2020 and estimated a country-specific lag time to the first infected cases in each country. To forecast accurately, it was essential to properly incorporate the population at risk for infection in each country and to avoid overestimating or underestimating the transmission rate because this could impact the model’s outcomes and ultimately misinform subsequent decisions regarding medication development and deployment. To account for non-uniform geographical distribution of cases within a country, the size of a susceptible population was initially estimated by mimicking the size and distribution of COVID-19 hotspots and was inflated every 15 days using an estimation of the inflation parameter for that country.

The asymptomatic population was included in our SEIR model by estimating the proportion of asymptomatic cases and assuming a daily infection rate of half that of symptomatic cases. This rate assumption was based on reports of reduced viral load in asymptomatic persons, a potential surrogate marker of the infection rate [23, 33, 34]. The infectious period of asymptomatic cases was fixed at 10 days, based on the observed viral load time course [23, 33]. These characteristics of asymptomatic cases were then further refined using sensitivity analysis. The super-spreaders were accounted for in our SEIR model in the same way asymptomatic cases were. However, both the proportion of super-spreaders in the population and the increase in their infection rate could be estimated in our model.

The effect of mitigation strategies on the RR (number of new cases per subject during the entire infectious period) was evaluated using the Oxford COVID-19 Government Response Tracker [32], which calculates a stringency index to score the strength of mitigations. This tool systematically collects country-specific policy responses to COVID-19, including indicators such as school closures and travel restrictions (Fig. 2). The value of an index on any given day is calculated as the average of nine sub-indices pertaining to individual policy indicators assigned a value between 0 and 100, where the stronger the mitigation, the higher the stringency index. In our model, the relationship between the stringency index and the daily infection rate was characterized using an Emax model from which a maximum infection rate could be estimated considering no mitigation (stringency index of 0), and then a decrease in the infection rate could be estimated depending on the stringency index, the magnitude of the decrease and the stringency index value that would correspond to 50% of the decrease.

**Figure 2.**
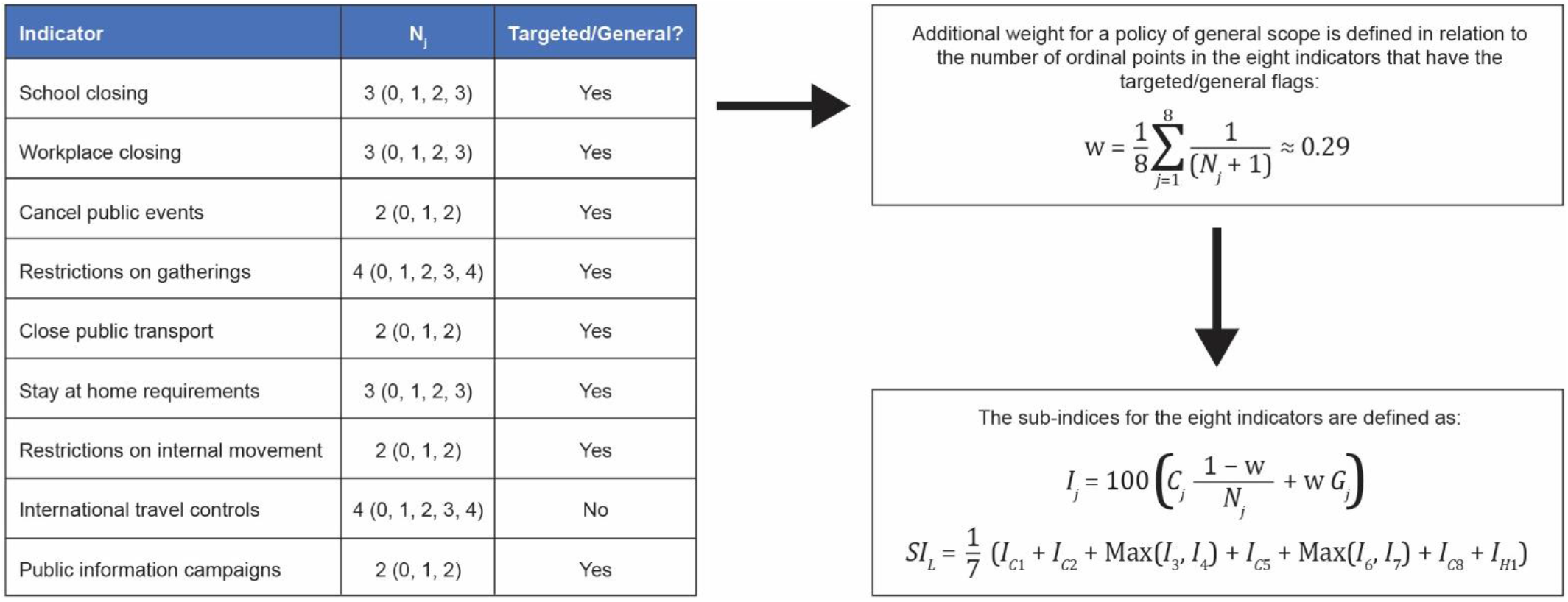
Stringency index for mitigation strength scoring. C_j_, ordinal value of the indicator; G_j_, general value; I_j_, subindex; N_j_, maximum value of the indicator.

We also assumed a similar relationship between the stringency index and the daily infection rate for symptomatic and asymptomatic cases, considering that different NPIs constituting the stringency index (e.g. school or public transport closing) would impact both infection rates. The policy most specific to symptomatic cases is to quarantine starting soon after the emergence of symptoms; we included this in the model as directly impacting the infectious period of symptomatic cases only.

### Between-country variability

Because our objective was to describe and forecast the number of cases in each country, we had to consider which parameters would be similar across countries (virus specific) and which parameters would vary between countries (country specific). Therefore, we included a certain amount of inter-country variability in our model to account for country-specific factors, among them potential differences in transmission resulting from cultural and environmental differences and differences in the way cases were reported. Similarly, the impact of mitigation strategies on the rate of transmission was considered country specific.

### Simulations

Our model was used to project the mid-to long-term expected number of cases in each country according to different scenarios. We fixed the stringency index at the latest reported value for each country at the cut-off date (25 October 2020) because it appeared to reflect the maximum sustainable mitigations countries could implement without jeopardizing economic factors. However, different scenarios could account for specific viral spreading according to the size of the target population (to mimic the occurrence of new clusters). We defined two possible scenarios. In scenario 1, which corresponds to low viral spread, the susceptible population was increased every 15 days using country-specific means of the stringency index values estimated during the period with strong travel limitations (mid-March to mid-May). Scenario 2 corresponds to high viral spread with a bi-weekly increase of the susceptible population implemented using country-specific means of the stringency index values estimated over the recent months with no travel restrictions (mid-August to mid-October).

We limited the simulation period to 2 months as we considered the effects of certain parameters (e.g. seasonality of virus transmission, effects of face mask wearing, test strategy, increase in reporting rate due to test deployment) sufficiently ambiguous to limit confidence in a longer simulation period.

## Results

### Tailored model for understanding virus spread characteristics per country

Parameter estimates are shown in Table 1. Across countries, our model estimated that it takes an average of 3.43 days after a person contracts the virus to become infectious and another 2.57 days before the onset of symptoms, resulting in an incubation period of almost 6 days. On average, 65.8% of all cases are estimated to be asymptomatic and 9.55% of all cases are estimated to be super-spreaders, with a transmission rate 2.11-fold higher than average.

**Table 1.** Population parameter estimates

| **Model parameter** | **Description** | **Value** | **Precision, %** | **Variability between countries, %** |
| --- | --- | --- | --- | --- |
| $\lambda$ | Time to becoming infectious after contracting virus, days | 3.43 | 0.187 | None |
| $\alpha$ | Time to development of symptoms after becoming infectious, days | 2.57 | 0.472 | None |
| kq | Time to starting quarantine after symptom onset, days | 1.00 | Fixed | None |
| Pra | Proportion of asymptomatic cases, % | 65.8 | 0.148 | None |
| Pss | Proportion of super-spreaders, % | 9.55 | 0.544 | None |
| $\beta_{ss}$ | Transmission rate increase in super-spreaders | 3.11 | 10.8 | 139 |
| lagD | Time to the first two infected persons, days | 23.1 | 5.48 | 71 |
| $\beta_{Max}$ | Maximum infection rate | 0.927 | 5.11 | 67 |
| $\beta_{min}$ | Minimum daily infection rate | 0.228 | 9.59 | 123 |
| S50_pop | Stringency index needed to reach 50% of the maximum effect on the infection rate | 35.3 | 9.33 | 117 |

The stringency index, derived from the Oxford COVID-19 Government Response Tracker [32], was used for scoring initiated mitigations. In our model, the maximum infection rate in the absence of specific policies (other than self-imposed quarantine in response to symptoms) was estimated as 0.927 cases per subject per day of symptoms, with a range between 0.63 and 1.41, depending on the country. Assuming a reduction in the infection rate of 50% for pre-symptomatic and asymptomatic cases, the average RR was computed as shown in Table 2. Based on this, a symptomatic person is predicted to infect a total of 2.12 people – approximately 1.1 during the pre-symptomatic period and 1.0 during the symptomatic period. An asymptomatic person is predicted to infect 5.83 people in total. Accounting for the proportion of asymptomatic cases estimated by the model, it can be derived that asymptomatic persons are responsible for 84% of new infections. Thus, the relationship between the stringency index and the daily infection rate could be estimated within the model; an example based on global level data is shown in Fig. 3. A stringency index value of 35.3% was estimated as required to result in a 50% decrease in daily infection rate with a range between 17.4% and 74.5%, depending on the country. Therefore, the stringency index must exceed 60% to result in the RR in symptomatic cases dropping below 1 (Fig. 3).

**Table 2.**
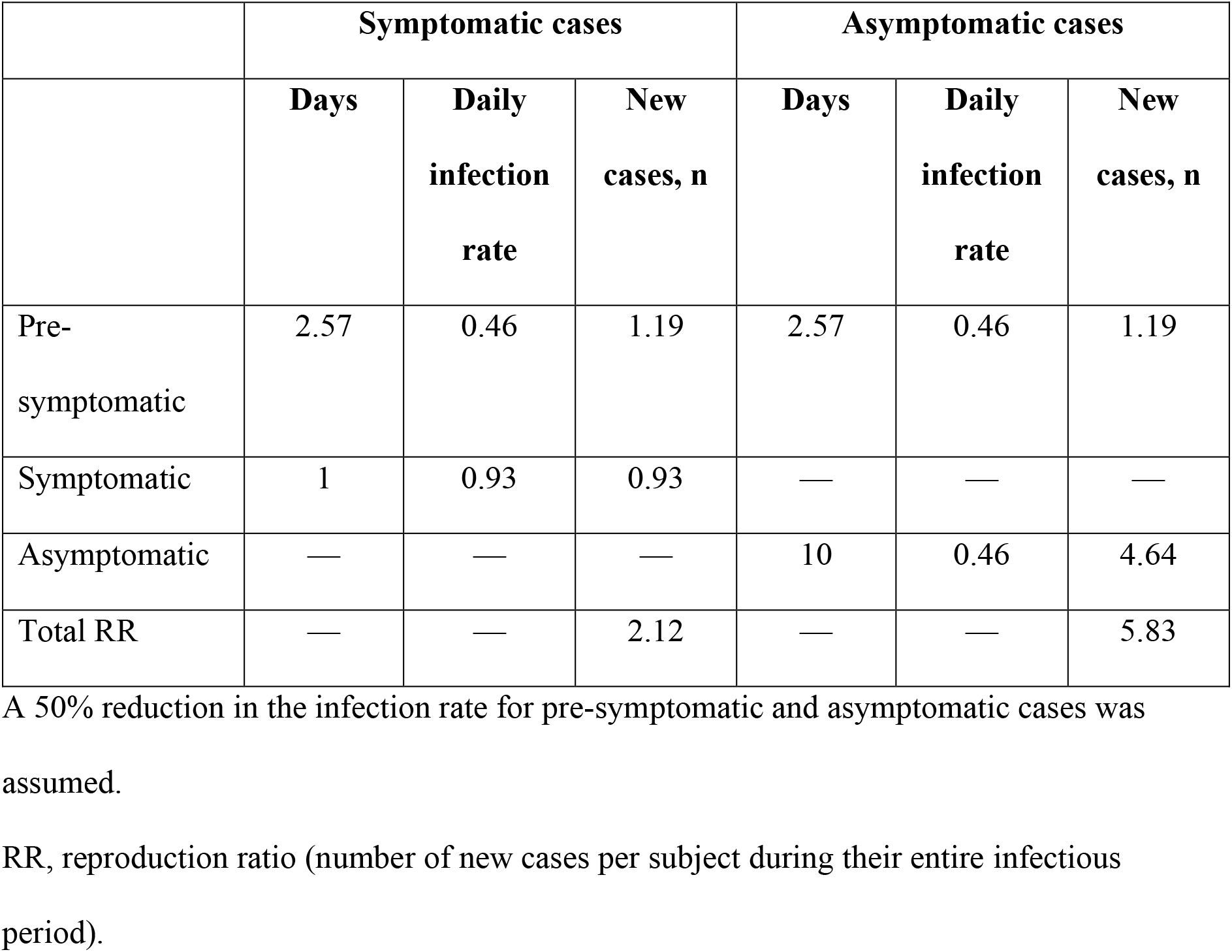
Estimated RR of symptomatic and asymptomatic cases

**Figure 3.**
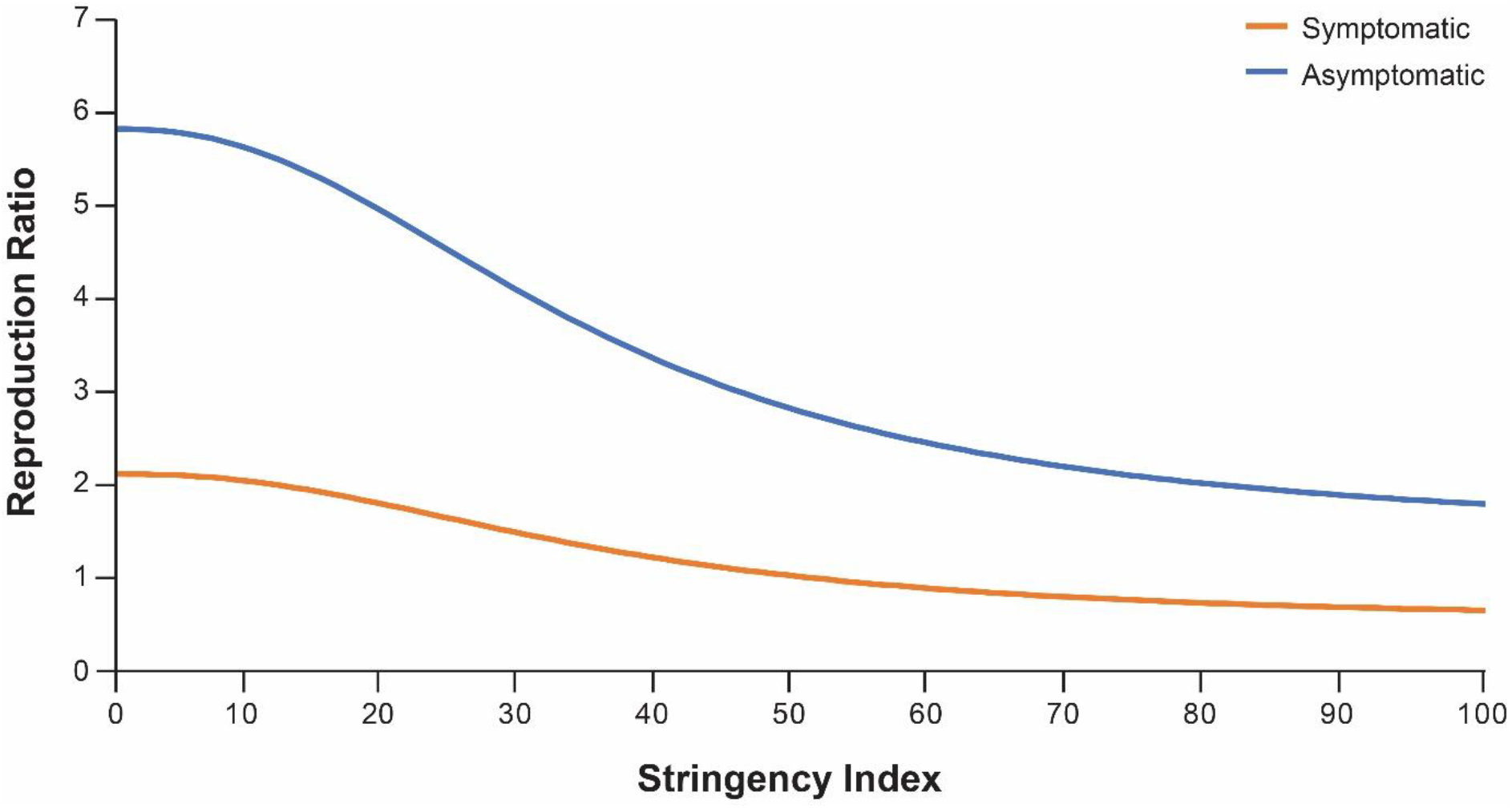
Relationship between the stringency index and the reproduction ratio.

Most of the parameters were well estimated. Data fitting to describe the observed cumulative cases are shown in Fig. 4 for representative countries and in the online supplementary appendix for all countries examined. Between-country differences observed in cumulative cases over time were well captured with the model and could be explained by changes in the stringency index and the potential geographic spread of the virus, which are the two time-dependent variables in the model.

**Figure 4.**
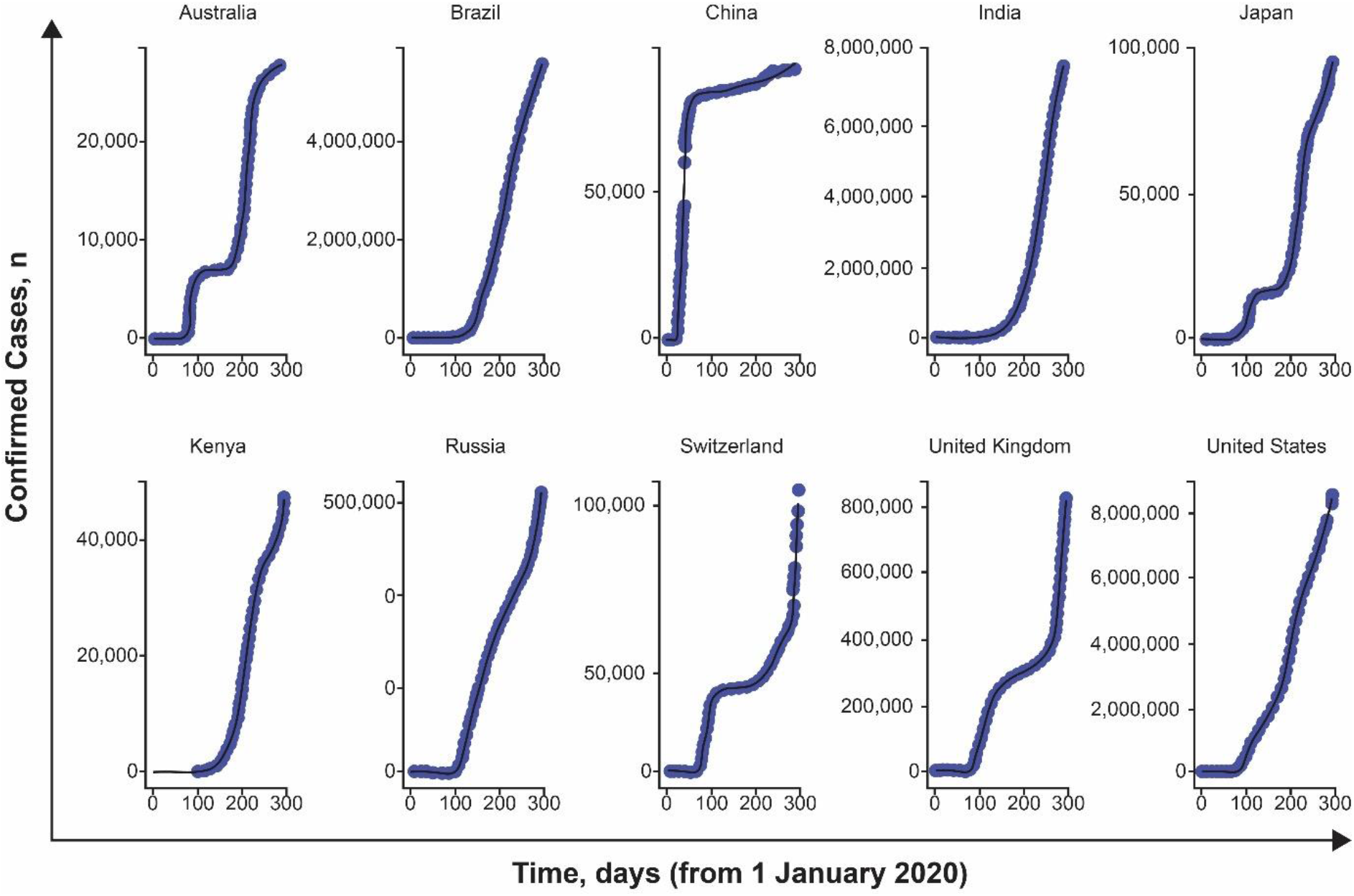
Data fitting for representative countries. Blue dots represent observed data; black lines represent model prediction. See online supplementary appendix for all countries.

### Supporting questions around clinical development and future supply

The simulation was also able to illustrate the projection of expected cases for the next 2 months (Fig. 5). These projections may help inform clinical operation considerations with regards to site location for new COVID-19 clinical trials and allow for companies to anticipate future demands for medications and prioritize supplies in territories with the highest current or future needs. For each of the scenarios tested, some countries, such as Russia and Peru, are likely to have more accelerated accumulations of cases than other counties, such as France and the United Kingdom, especially as travel restrictions are put in place in countries with slower accumulations of cases. The ability to predict where this acceleration will occur may allow for a more appropriate selection of clinical trial sites for new COVID-19 medications and may increase patient participation in these clinical trials. This, in turn, will ensure that medications that are safe and effective can be rapidly distributed to the patients who need them. Such simulation results, if regularly revised, could also allow for increased availability of medications where the need is likely to be high while also identifying countries that may be less relevant targets.

**Figure 5.**
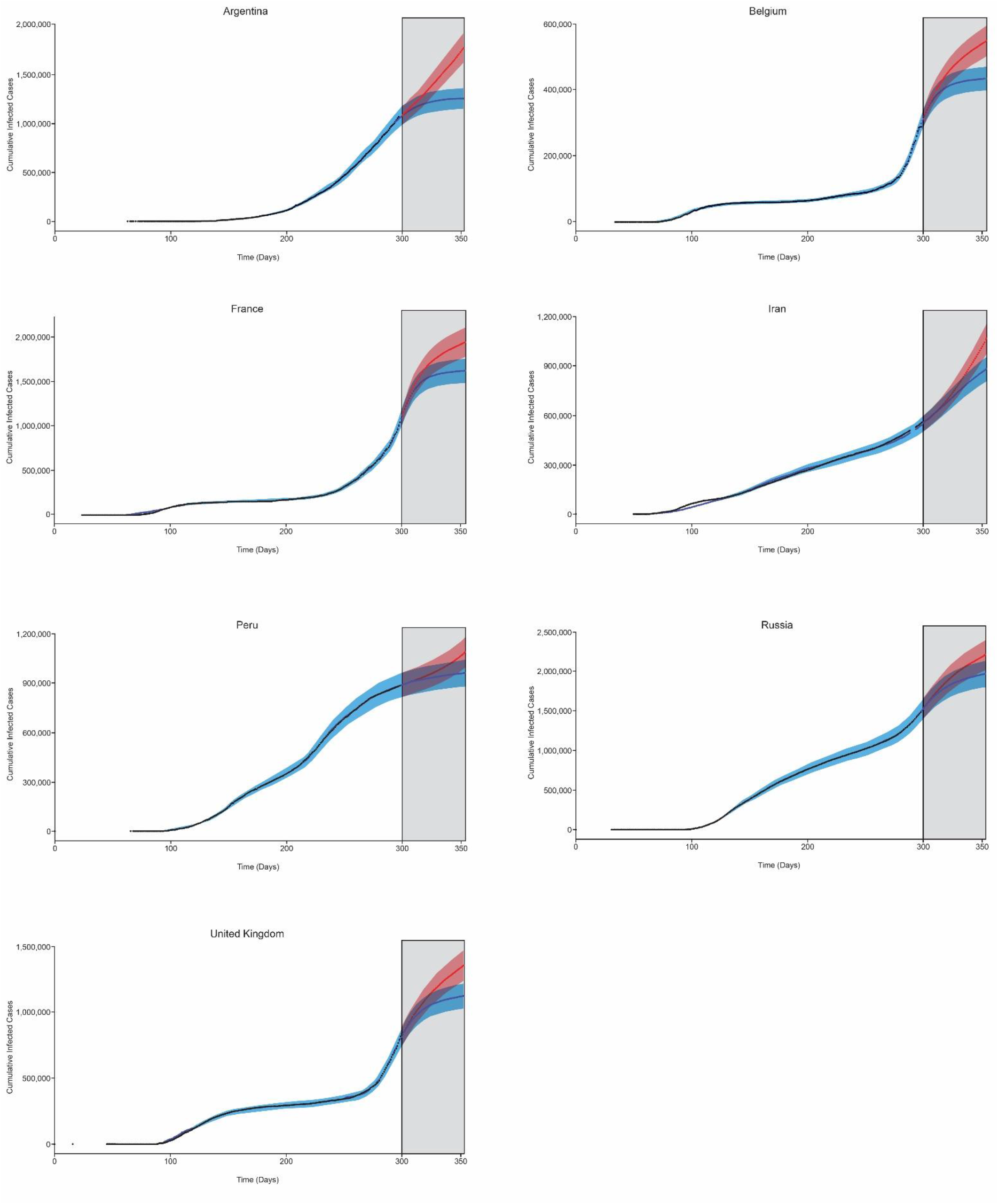
Simulation results by country. Black dots indicate observed data; blue shaded area indicates simulated time course for each scenario with low viral spreading; red shaded area indicates simulated time course for each scenario with high viral spreading; grey rectangles highlight the forecasted period.

## Discussion

The objective of this analysis was to tailor a transmission model to address the current issues faced by pharmaceutical companies, governments and multi-national organizations: how to best identify countries that can facilitate faster and more efficient development of potential COVID-19 therapies, with the ultimate goal of getting potentially life-saving medications to those in need. The model presented here has the potential to help support the selection of clinical trial sites, the initiation of mitigation strategies and the distribution of COVID-19 diagnostics, treatments and vaccines.

This modelling study of COVID-19 transmission, based on total global cases reported as of 25 October 2020, demonstrated disease model parameters consistent with those that have been reported in the literature, such as an incubation period of 6 days [35, 36][37]. In addition, our model was able to estimate similar rates of asymptomatic cases and proportions of super-spreaders across all countries. The estimated proportion of asymptomatic cases (66%) is very close to other reported values in the literature [15, 23, 38]. In addition, the estimated proportion of super-spreaders identified here (almost 10%) is similar to the expected proportion identified in other studies [39]. According to their daily infection rate, super-spreaders are expected to infect 3.11 times the number of people normal spreaders infect. In the absence of mitigation, this would lead to one symptomatic super-spreader infecting approximately 6.6 people and one asymptomatic super-spreader infecting almost 11 people, highlighting the large contribution this small proportion of people can make to virus transmission, which is consistent with previous reports [40]. Furthermore, our model confirms that most new cases result from asymptomatic transmission, occurring either during the pre-symptomatic period (from persons who later become symptomatic) or from asymptomatic persons [27, 28]. In general, these data highlight the importance of incorporating asymptomatic persons in transmission models to obtain more accurate projections of future cases.

To better describe country-specific data, identify which countries are likely to experience large numbers of emerging cases and help inform decisions surrounding future clinical trial locations, our strategy incorporated inter-country variability into the model. This variability was considered for parameters that may differ, depending on social, cultural and societal factors (such as the rate of infection and the strength of NPIs), allowing for appropriate country-specific forecasts. These parameters could indirectly account for the number of contacts each person may have (e.g. household context, place of work) and the level of adherence to NPIs, with an effect that could differ between countries despite a similar stringency index. Notably, the amplitude of the inter-country variability was large, suggesting large heterogeneity between countries.

With this model, we were able to simulate the expected number of COVID-19 cases over time using different scenarios of mitigation. Our model can be used to project the future profile of the pandemic based on current data to investigate the effect of mitigation specific to each country. It also allows for the simulation of potential new clusters though a country-specific stringency index (i.e. last stringency index carried forward in the simulation), accounting for an increase in the target population over time based on country-level estimates. Together these data can provide important information for the identification of promising new sites for COVID-19 clinical trials and may help support global supply chain networks by identifying potential supply-and-demand challenges arising in different countries during the pandemic.

Our model does have certain limitations. First, it was based on observed infected cases. Given that some patients are asymptomatic or show only mild symptoms, however, it is likely that the true number of cases is higher than captured here. In addition, changes in the reporting rate because of local testing policies and potential seasonality differences are not incorporated or investigated in this model. As data on these factors are gathered over time, it may be possible to integrate them into our model to support future forecasting. Nevertheless, it is worth noting that this model has remained relatively stable since September 2020, with bi-weekly updates only minimally impacting the forecasting results. Finally, the model does not account for the availability of a vaccine. When an efficacious vaccine is widely available, our model must be adjusted to account for the reduced size of the susceptible population and the limited possibility of transmission. As with all predictive models, additional data on each of these factors will improve our understanding of SARS-CoV-2 transmission; integrating these data into our model can support more accurate forecasting in the future.

In summary, our model can support and inform the development of clinical trials and the supply and distribution of future medications. By updating and adjusting the model as new data are received, our model could potentially inform longer-term considerations as well. Finally, our model also represents a possible framework for describing transmission characteristics of other diseases, estimating viral spread and refining country-specific estimates of disease impact.

## Data Availability

Qualified researchers may request access to individual patient level data through the clinical study data request platform (https://vivli.org/). Further details on Roche's criteria for eligible studies are available here (https://vivli.org/members/ourmembers/). For further details on Roche's Global Policy on the Sharing of Clinical Information and how to request access to related clinical study documents, see here (https://www.roche.com/research_and_development/who_we_are_how_we_work/clinical_trials/our_commitment_to_data_sharing.htm).

https://vivli.org/members/ourmembers/

https://www.roche.com/research_and_development/who_we_are_how_we_work/clinical_trials/our_commitment_to_data_sharing.htm

## Acknowledgements

Third-party editorial assistance was provided by Sara Duggan, PhD, and Stacie Dilks, PhD, of ApotheCom and was funded by F. Hoffmann-La Roche Ltd.

## Author contributions and degree of contribution

Conceptualization and writing review and editing: PB, JEC, BC, SF and AH; data curation: ML; formal analysis: ALD; investigation: PB; methodology: ALD, PB and JEC; resources: ML; software: JL; supervision: JEC.

## Financial support

This study was funded by F. Hoffmann-La Roche Ltd.

## Conflict of interest

ALD, JL and ML are employees of Roche Pharmaceuticals, Inc., and receive salary and stock options. BC is an employee of Roche Products Ltd and receives salary and stock options. ACH, PB, SF and JEC are employees of F. Hoffmann-La Roche Ltd and receive salary and stock options.

## Notes

### Author Declarations

Real-world epidemiological data were obtained from the Johns Hopkins University COVID-19 Center for Systems Science and Engineering COVID-19 Dashboard on 31 August 2020. This includes data from national and state government health departments and local media reports. In addition, country-level mitigation data from the Coronavirus Government Response Tracker, collected and validated by Oxford University, were used to investigate the potential mitigation impact of NPIs on the transmission of SARS-CoV-2, with the objective of building country-specific quantitative relationships between NPIs and transmission model parameters.

